# Using Random Forests with Asymmetric Costs to Predict Hospital Readmissions

**DOI:** 10.1101/2021.03.15.21253416

**Authors:** Justin Bleich, Brian Cole, Adam Kapelner, Charles A. Baillie, Rohit Gupta, Asaf Hanish, Erwin Calgua, Craig A. Umscheid, Richard Berk

## Abstract

**Background:** Sufficiently accurate predictions of hospital readmissions are necessary for the allocation of scare clinical resources to reduce preventable readmissions. We describe the use of a data-driven approach that relies on machine learning algorithms to predict readmission at the time of discharge.

**Methods:** We employ random forests to clinical and administrative electronic health record data available from a cohort of 103,688 patients discharged from the acute inpatient settings of the University of Pennsylvania Health System between June 25th, 2011 and June 30th, 2013. We predict both 30-day all-cause readmissions and 7-day unplanned readmissions using only predictors available by the time of discharge. Using oversampling and undersampling of the different outcome classes of readmission and no readmission, we incorporate into our models the asymmetric costs of a false negative relative to a false positive from the perspective of a hospital. We calculate variable importance scores for included predictors. Our approach was derived and validated using split-sample internal validation.

**Results:** We developed a machine learning-based model using random forests with a 5:1 relative cost ratio for 30-day all-cause readmissions that achieves a sensitivity of 65% and specificity of 71% on validation data, as well as a random forests model with a 20:1 cost ratio for 7-day unplanned readmissions that achieves a sensitivity of 62% and specificity of 66% on validation data. Prior health system utilization, clinical discharging service, and vital sign information were most predictive of readmissions.

**Conclusion:** By modeling the complex relationships between many predictor variables and readmission data for a large health system, we demonstrate successful predictive models that can be used upon discharge to flag patients at high risk of readmission.

## 1 Background

Hospital readmissions (the return of a patient to the hospital within some specified time after discharge) often reflect poor quality care, and can result in significant patient distress and high health care costs [1–6]. Thus, reduction in hospital readmissions is a public health priority. In the United States, the Centers for Medicare & Medicaid Services (CMS) penalizes health care providers with high rates of 30-day all-cause readmissions [7, 8]. Therefore many hospitals have an interest in accurately predicting patients’ risk of readmission in order to target their limited resources to prevent them [4, 9–11].

Prior approaches to predict readmissions using regression models and clinical and administrative data have had poor to modest predictive power [4, 7, 12–15]. Additionally, preexisting models are often disease-specific with potentially limited generalizability [2, 6]. Further, all of these approaches have used manual regression modeling to select risk variables [2, 8], a process that is particularly cumbersome when it is estimated that an acutely ill inpatient generates a median of 1,348 individual data points per patient per day[16], ranging from laboratory values to free text data [15]. This has created a need for novel approaches to prediction, such as the application of machine learning algorithms and natural language processing, which take advantage of “big data” available from electronic health records (EHR).

EHR data present the opportunity to leverage machine learning algorithms and data mining techniques to uncover previously unknown and often complicated patterns and associations that can be used to improve quality of care and inform clinical decision-making in real-time. The advent of affordable data storage and analysis technologies at the health system level has opened new frontiers in institutional evidence-based practice, including the application of predictive modeling techniques such as machine learning to identify patients at high risk of adverse events such as readmission.

Machine learning approaches have been successfully employed to predict adverse clinical outcomes using high-dimensional datasets such as EHR data, often significantly outperforming traditional regression approaches [17, 18]. In particular, the random forests (`RF`) algorithm [19] was developed to provide highly generalizable predictions based on an ensemble of tree-based classifiers that are individually derived from random subsamples of a training dataset. `RF` has been successfully employed as a predictive classifier across diverse application domains such as criminology [20], ecology [21], and bioinformatics [22]. Additionally, `RF` classifiers have shown the strongest performance in a large empirical study of 179 statistical models and machine learning algorithms [23].

A perceived limitation on the predictive performance of `RF` (and similar classifiers including logistic regression) appears when one class (or outcome) occurs less frequently than another in a dataset, as is the case with hospital readmissions, which occur relatively infrequently. The perceived limitation arises because the algorithm is designed to favor predicting the majority class (or most frequent outcome) in order to minimize overall misclassification error [22]. However, minimization of overall misclassification error implicitly assumes that the different types of misclassification errors (i.e. false positives and false negatives) have equal costs. Hospitals, on the other hand, face asymmetric costs for the different misclassification errors related to readmissions. In other words, falsely misclassifying patients as low risk and thus foregoing opportunities to prevent readmissions is likely more costly given existing financial penalties than falsely misclassifying patients as high risk, and providing transitional care services when they may not have been needed. By default, `RF` assumes equal costs and optimizes the misclassification rate under the assumption of these equal costs. In this scenario, where the costs of the different misclassification errors are assumed equal and the event rate for a particular outcome is low, the algorithm can achieve the lowest total misclassification rate by predicting no readmissions for all hospital discharges. These limitations of `RF` can be overcome by modifying the `RF` procedure to incorporate the appropriate asymmetric costs during model construction. We offer such an approach herein. Additionally, we argue that the mechanism by which `RF` incorporates asymmetric costs is favorable compared to the mechanisms utilized by alternative algorithms.

We additionally demonstrate models that could be deployed at time of discharge, thereby allowing the health system to direct transitional care interventions and intensive discharge planning to patients at high risk of readmissions. Implementation of these models provides the potential for avoiding adverse outcomes that threaten patient safety and generate high costs for the health care system.

### 1.1 Objective

In this study, we describe the development and resulting performance of `RF` modeling approaches to predict both 30-day all-cause and 7-day unplanned readmissions. We utilize two key methods in our approach. First, we restrict our set of patient variables to only employ clinical and administrative data available to the health system by the time of a patient discharge, allowing for real-time prediction. Second, we construct cost-sensitive models that can accurately account for the relative rarity of readmission events and the asymmetric costs associated with misclassification errors related to readmission events.

## 2 Methods

### 2.1 Setting

The University of Pennsylvania Health System (UPHS) includes three hospitals with over 1,500 beds and more than 70,000 annual admissions. We obtained data on 103,688 adults discharged from the acute inpatient settings of UPHS between June 25th, 2011 and June 30th, 2013. The unit of analysis is each unique hospital visit. Thus, individual patients can have multiple visits within the data set, and for each admission, a prediction is generated. The study received expedited approval and a Health Insurance Portability and Accountability Act (HIPAA) waiver from the University of Pennsylvania Institutional Review Board.

### 2.2 Outcome Variables

Our primary outcome of interest was all-cause hospital readmissions to any of the hospitals within the healthcare system within 30 days of discharge. While this outcome is commonly used both in the literature and as a quality metric, a shorter window for readmissions may offer a more meaningful measure of hospital quality [24]. As a consequence our health system adopted 7-day unplanned readmissions as an internal quality metric. Seven-day unplanned readmissions as defined by the health system were thus used as a secondary outcome measure for this study. Unplanned readmissions excluded elective or scheduled readmissions, as well as admissions for ventricular assist device placement, transcatheter aortic valve replacement, transplants, and deliveries [25].

### 2.3 Predictor Variables

We examined 188 variables (e.g. predictors) for each hospital admission. Classes of predictors include information on demographics, previous admissions statistics, lab test values and vital sign recordings. For predictors with more than two values during a given hospital encounter (e.g. lab test and vital sign information), we retained only the first and last values recorded as well as the times of the recording. We excluded predictors that were not available by the time of discharge, such as procedure and diagnostic codes for that admission.

For quantitative predictors with large proportions of missing data such as laboratory tests, we transformed quantitative values into categorical predictors consisting of deciles and added an additional level for missing values. For these recoded quantitative predictors, we additionally relabeled extreme values (defined as being in the top or bottom 0.5 percentile of a given predictor’s distribution) to a level denoted as “extreme”.

### 2.4 Random Forests for Prediction

`RF` are capable of achieving excellent predictive performance by relying on an ensemble of classification trees. Classification trees are a nonparametric statistical method that are capable of capturing nonlinearities and interaction effects in data. In a classification tree, all data begins in a root node and the tree searches for binary partitions of the data that result in the most homogeneity within the two resulting splits. From the two new splits, further splits are similarly computed in a recursive fashion until some stopping rule is achieved. Observations rest in a “terminal node” of the tree in which classifications can be assigned by computing the majority class within the node. For new observations, predictions can be made by assigning the majority class of the terminal node in which the new observation lands.

As standalone prediction models, classification trees have relatively weak predictive accuracy [26]. The `RF` algorithm improves upon a single tree by building an ensemble of many classification trees via “bagging”. For each tree, a bootstrap sample is drawn from the original data and the tree is constructed on this data. Additionally, only a random subset of predictors is searched at each partition of the classification tree rather than the full set of all predictors. This random predictor selection reduces correlation across trees and further bolsters predictive accuracy.

Fitted values for the training data are computed using “out-of-bag” (OOB) estimates. First, for each tree in which an observation was not included in the bootstrap sample, a prediction is obtained. Then, the OOB estimate for a given observation is the majority vote across all trees where the observation did not participate in growing the tree. Since the OOB estimate for each observation is computed only on trees the observation did not help construct, OOB estimates are good approximations of predictive performance on future data and can be used to guide model construction. Finally, predictions on future data can be computed by obtaining the vote for each tree and then taking the majority vote across the individual tree votes.

To construct our `RF` models, we used the `randomForest R` package [27]. We chose default parameters for the number of trees (*ntree* = 500) and the number of predictors selected at each split 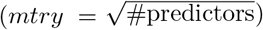. We alter the *sampsize* parameter in order to introduce asymmetric costs into the model. The proper amount of sampling is specific to each data set’s composition and the desired cost ratio. Further details are provided in the subsequent section.

### 2.5 Random Forests with Asymmetric Costs

As previously discussed, the cost of an incorrect prediction of readmission (false positive) is not likely to be equal to that of an incorrect prediction of no readmission (false negative). This cost asymmetry may arise from resource considerations and constraints, such as a limited staff to employ readmission reduction interventions, as well as financial penalties associated with hospital readmissions. Here, we consider failing to identify a readmission to be relatively more costly. For the 30-day model, we consider a failure to identify a readmission (i.e. a false negative) as 5 times more costly than incorrectly predicting a readmission for a patient who is not readmitted (i.e. a false positive), generating a 5:1 cost ratio. Given the relative rarity of 7-day unplanned readmissions, there must be a substantial cost incurred by the health system for failing to identify patients with this type of readmission, else these cases are not worth identifying from a cost minimization perspective. Thus for the 7-day unplanned readmissions, we consider a cost ratio of 20:1. Note that these cost ratios are meant to be illustrative and should be appropriately determined based on each hospital’s unique circumstances.

The `RF` must be properly tuned to reflect the asymmetric costs. We incorporate these cost ratios into the model by sampling about 2/3 of the cases with no readmission and oversampling or undersampling the cases resulting in readmission until the desired empirical cost ratio is achieved on the OOB estimates [20]. The resampling is computed independently for each tree in the forest and not globally on the entire dataset. Additionally, using 2/3 of the no readmission cases in each tree balances the training of the model on the rare outcome with leaving enough OOB observations for evaluation. The approach is similar in spirit to the balanced `RF` approach [28], but we do not necessarily use equal sampling sizes for the classes. By relying on this resampling approach, each tree in the `RF` is constructed with the cost asymmetry taken into account. Not only will the predicted classes reflect this asymmetry, but any additional model output, such as variable importance scores, will as well. Additionally, since each tree draws a unique stratified bootstrap sample based on the desired sampling size values for each class (given by the *sampsize* parameter), different observations appear in each tree. Hence, even when a particular class is undersampled, all observations are likely to appear in at least some trees and can contribute to the overall ensemble. Rebalancing the dataset via undersampling before deploying `RF`, on the other hand, would result in a loss of data and potential predictive information [28].

Note that asymmetric costs can also be accounted for by altering the threshold on the voting rule for classification. For example, rather than using majority vote, 2/3 of the trees might be required to classify a case as readmission in order to assign a final label of readmission. A major drawback of this approach is that asymmetric costs are only incorporated after the model has been fully constructed. The asymmetric costs are not accounted for in the tree-growing process [29].

### 2.6 Random Forests versus Alternative Algorithms

Compared to a parametric logistic regression model, our machine learning approach offers a number of significant advantages. First, the machine learning algorithms are able to discover rich relationships between predictors and the response automatically, such as predictor transformations or high-order interaction effects. While capturing these relationships is in principle possible for logistic regression, the challenge is that the researcher must specify such effects *a priori*. Second, machine learning procedures can easily handle more predictors than the number of observations. Finally, machine learning algorithms are purposefully designed to maximize out-of-sample forecasting accuracy rather than maximize in-sample fit as in logistic regression [20, 29].

Additionally, `RF` maintains extra advantages over competitor machine learning algorithms and regression approaches in asymmetric cost settings. Logistic regression as well as a number of alternate machine learning algorithms such as stochastic gradient boosting [30] and Bayesian additive regression trees Chipman et al. [31] can only incorporate asymmetric costs by altering the threshold on their respective probability estimates. This implies that information about the model fit (such as regression coefficients or tree-based variable importance scores) does not reflect the asymmetric costs as the costs are applied only to the model output and not the model itself. An advantage of the resampling approach used in `RF` is that the asymmetric costs flow through the entire fitting procedure, implying *all* output associated with the model reflects stakeholder preferences, and not just the predicted classes.

While `RF` is a robust and effective choice for prediction under asymmetric costs, the major weakness of `RF` is the difficulty in interpreting the model. By combining trees, it is difficult to assess which predictors drive the response as well as the functional form of such relationships. Traditional statistical inference, such as hypothesis tests, is also generally unfeasible. Additionally, random forests tend to perform poorly on small datasets as the trees are unable to grow deep enough to successfully model complex data relationships [20].

### 2.7 Variable Selection

In theory, a health system could construct a `RF` approach using the full set of predictors available by the time of discharge and allow the model to dynamically determine which predictors are most useful. From an implementation perspective, however, it can be difficult to always collect such a complete set of information for each patient in real-time. Therefore, we strive to develop models using a smaller subset of the predictors without sacrificing predictive performance. This is achieved via a backward selection that iteratively removes predictors with the lowest “variable importance scores”[19, 32]. For each predictor, the variable importance score we use represents the average increase in prediction error for a given outcome class when the predictor of interest is not allowed to contribute to the current model. Hence, predictors with higher scores (and thereby larger implied performance losses) are more important. Our elimination procedure continues removing predictors until the cost-sensitive performance begins to degrade when compared to performance when using the full set of predictors available. No stopping criterion was employed, but rather the cost-sensitive performance results were manually inspected to determine the final set of predictors. We utilized the variable importance scores computed within the randomForest package and describe these scores in detail in Section 3.2.

### 2.8 Model Evaluation

In any machine learning application, it is of utmost importance to have insight into how this model will perform on future data before actual implementation into the production environment. It is possible to obtain an accurate assessment of future performance, but proper procedures must be followed. The variable selection procedure and model construction must occur on one subset of the data (the training data) and then evaluated on a different subset of the data (the validation data). The validation data should not be used for model construction, otherwise any estimates of the models future performance using this test data would be invalid. Thus, we performed variable selection and model development on a 75% random subset of the data, reserving the remaining 25% to validate model performance. Given the large number of observations, we anticipate that results from split sample validation should be stable. Additionally, note that under this scheme, we are implicitly assuming stationarity with respect to time across the sample period. That is, the relationship between the predictors and readmission risk does not change over the time period we examine.

## 3 Results

### 3.1 Descriptive Characteristics

Table 1 provides a summary of the cohort of patients used in this study.

**Table 1:** Study cohort characteristics.

| Characteristic | Value |
| --- | --- |
| Mean age, years (SD) | 58.5 (17.2) |
| Female, % | 50.1 |
| Race/Ethnicity, % |  |
| White | 56.2 |
| Black | 36.7 |
| Other | 7.1 |
| Median length of stay (IQR), days | 3.7 (2.6, 6.5) |
| Discharging service, % |  |
| Medicine | 56.2 |
| Surgery | 22.6 |
| Mixed/Other | 25.8 |
| Top 5 primary discharge diagnoses, % |  |
| Circulatory system diseases | 22.2 |
| Neoplasm | 13.4 |
| Injury and poisoning | 13.3 |
| Digestive system diseases | 9.7 |
| Musculoskeletal system diseases | 9.4 |

Table 2 shows the incidence of 30-day all-cause and 7-day unplanned readmissions. Here we characterize all clinical services into either one of Medicine or Surgery. Overall, admissions to medical units result in slightly higher rates of readmissions than those to surgical units.

**Table 2:** Incidence rates for 30-day all cause and 7-day unplanned readmissions.

| Readmission Window | Medicine | Surgery | Overall |
| --- | --- | --- | --- |
| 7-day | 4.7% | 3.4% | 4.2% |
| 30-day | 13.8% | 8.1% | 11.3% |

We observed substantial heterogeneity of readmission rates when different services are compared. For example, categorizing individual clinical services into either Medicine or Surgery services, we observe that the Hematology/Oncology service attains the highest overall readmission rate (24.7% of visits result in 30-day all-cause readmissions and 9.1% of visits result in 7-day unplanned readmissions), and that the Transplant service had the highest readmission rate among surgical services (22.1% of visits result in 30-day all-cause readmissions and 9.9% of visits result in 7-day unplanned readmissions). These results demonstrate the importance of hospital service in prediction of readmission risk.

### 3.2 30-Day All-cause Readmissions Random Forests Model

We used OOB confusion matrix-derived performance metrics to achieve a sampling scheme that resulted in a 5:1 cost ratio as defined by the ratio of false positives to false negatives (Figure 1). The cost ratio achieved from OOB estimates was 4.97 (19,794 false positives divided by 3,977 false negatives), which corresponded to the 5:1 assumed stakeholder cost ratio preferences. Since there are about five times more false positives than false negatives, but each false negative is five times more costly, the overall cost of the prediction errors are roughly balanced.

**Figure 1:**
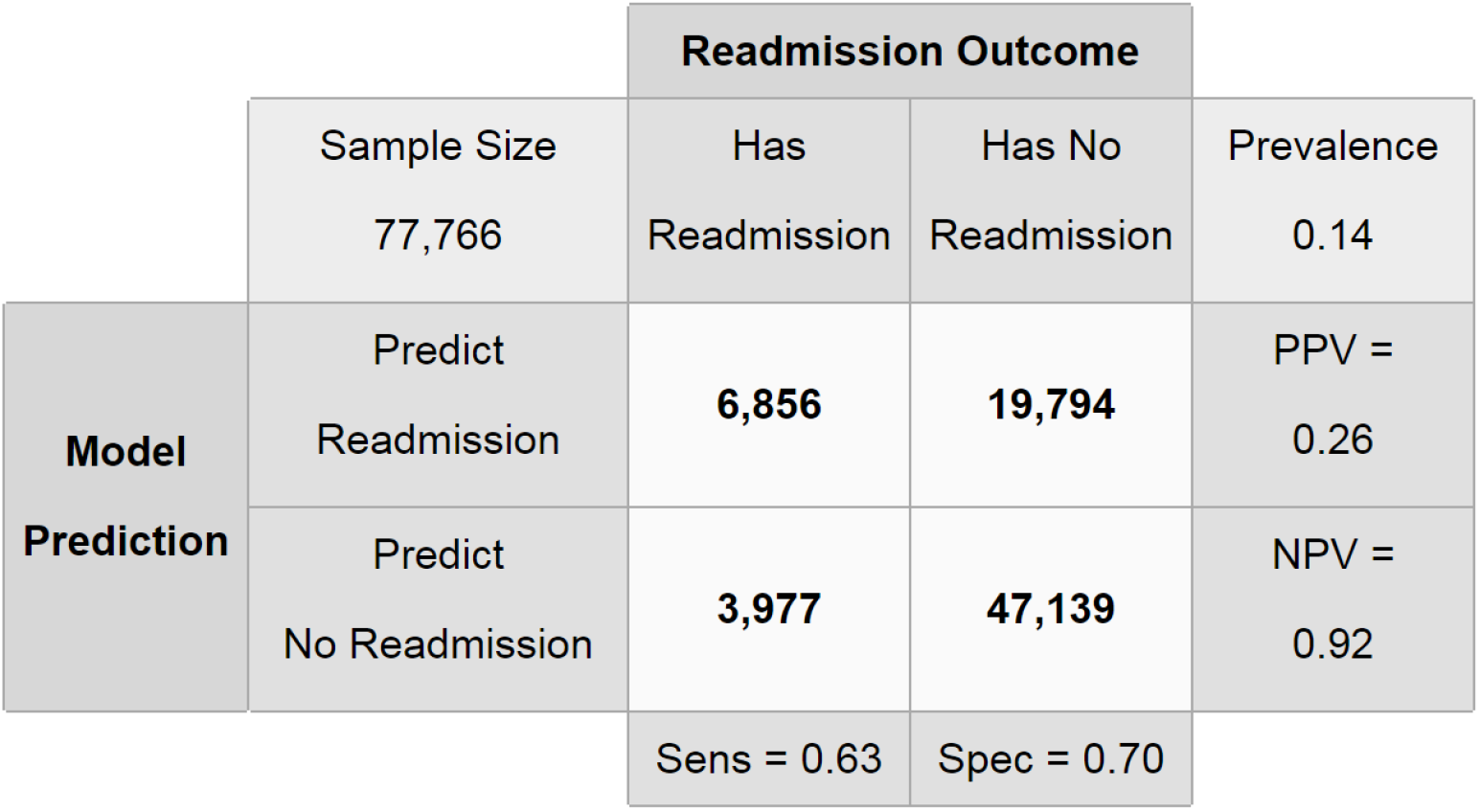
`RF` confusion matrix for 30-day all-cause readmissions on training data (OOB) using 5:1 cost ratio.

Performance conditional on our given cost ratio is examined via the row and column proportions in the confusion matrix. The column conditional proportions provide sensitivity and specificity measures, given as 63.3% and 70.4%, respectively. The two main rows of the confusion matrix give the positive and negative predictive values, given as 25.7% and 92.2% respectively.

The resulting model’s performance was then evaluated on the 25% validation data, where we observed a cost-ratio of 5.01 (6,528 false positives divided by 1,303 false negatives), demonstrating strong concordance with the OOB values from the training data subset and suggesting that the approach performs well on the validation data with respect to stakeholder preferences (Figure 2). Similar to the OOB performance on the training data, the model achieved a sensitivity of 64.7% and a specificity of 70.6%, and positive and negative predictive values of 26.8% and 92.3% respectively.

**Figure 2:**
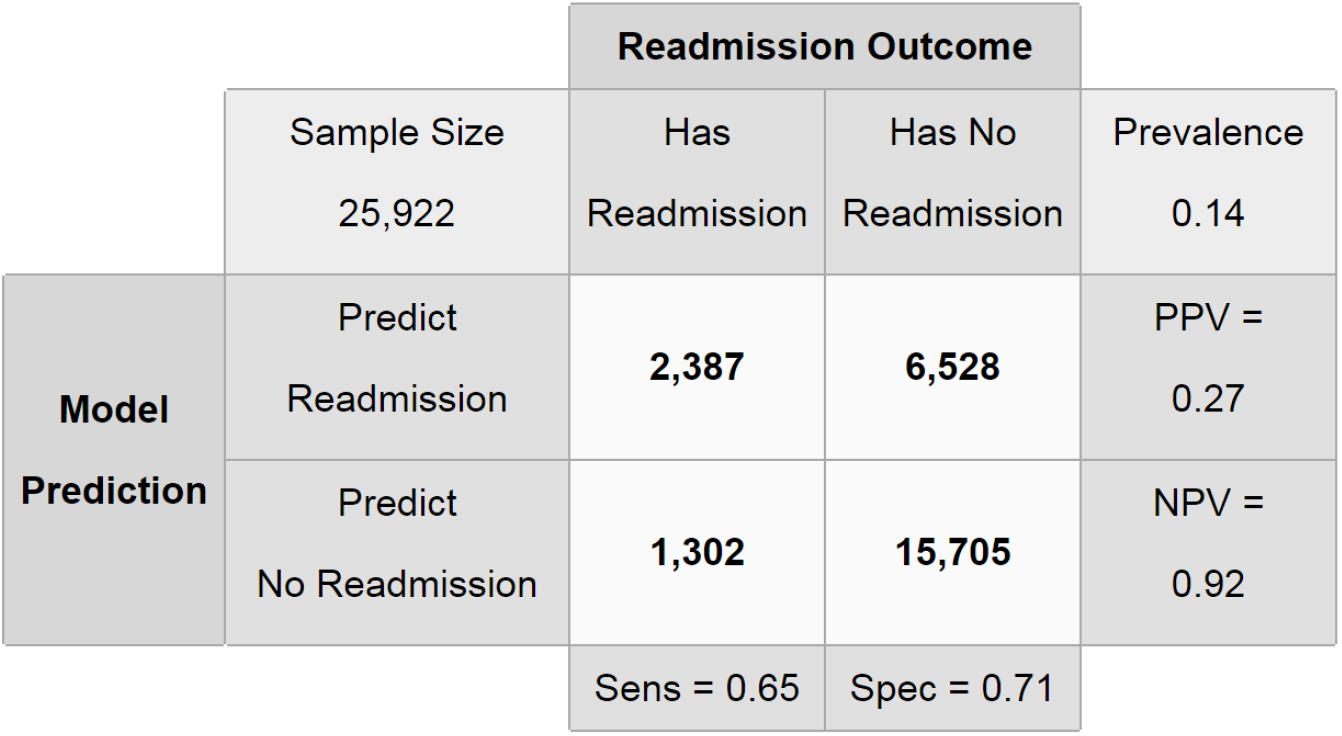
Random forests confusion matrix for 30-day all-cause readmissions on validation data using 5:1 cost ratio.

Given the performance of the model, one may inquire as to which predictors are contributing to the black-box fit constructed by the `RF`. The variable importance scores computed by `RF` can provide such insights, and the variable importance plot for the 5:1 30-day all-cause readmission model is shown in Figure 3. Recall that this score is to be interpreted as the increase in prediction error assuming that the predictor is not allowed to contribute to the current model. Hence, these scores are conditional on the existing model and do not apply to a model refit with a given predictor excluded. For example, Figure 3 indicates that if the predictor representing the total count of previous admissions is not allowed to contribute to the current model, predictive accuracy for the outcome “readmission” would decrease by about 2.0%. It is important to note that for variables further down the plot, such as the first heart rate value, despite the variable importance value being near 0, this does not necessarily imply that the variable has no predictive power. Variables in `RF` models are intricately tied together through the tree construction process and singly weak predictors can have large aggregate contributions. The plot only considers contributions to predictive accuracy one predictor at a time.

**Figure 3:**
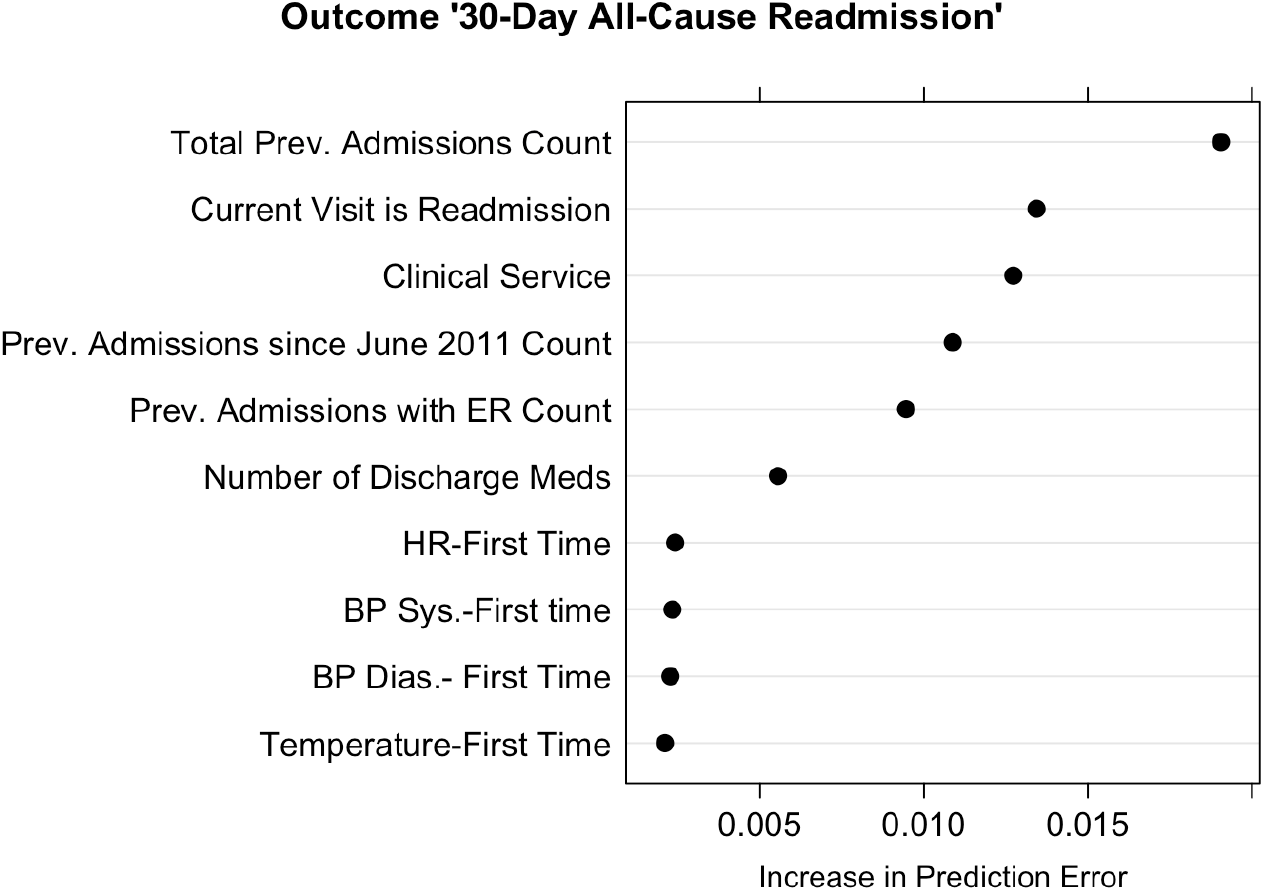
Variable importance plot for 30-day all-cause readmissions `RF` model. The top 10 variables for the class “readmission” are shown.

We included 39 predictors in our selected model, and the top 10 predictors are included in Figure 3. Prior health system utilization, the clinical service at discharge, and vital signs were among the most important predictors. For a full illustration of all variables’ importances, see the Appendix.

### 3.3 7-Day Unplanned Readmissions Random Forests Model

To provide additional insight into preventable readmissions, we focused on 7-day unplanned readmissions that were identified as unplanned by ICD-9 and DRG codes. We used a cost ratio of 20:1, reflective of the decreased prevalence of unplanned 7-day readmissions. The training model had a sensitivity of 61.2%, a specificity of 65.3%, and positive and negative predictive values of 7.1% and 97.4% respectively (Figure 4). The results for the validation data were similar (Figure 5). We observed an empirical cost ratio of 19.5, in accordance with the target cost ratio of 20:1. To assess the impact of predictors on unplanned 7-day unplanned readmissions, we analyzed the importance of the top 10 predictors as done for the 30-day all-cause model (Figure 6). The most important predictors were similar for the two readmission outcome timeframes, except that the serum sodium value at discharge appears in the top predictors of 7-day unplanned readmissions and blood pressure measurements do not. For a full illustration of all variables’ importances, see the Appendix.

**Figure 4:**
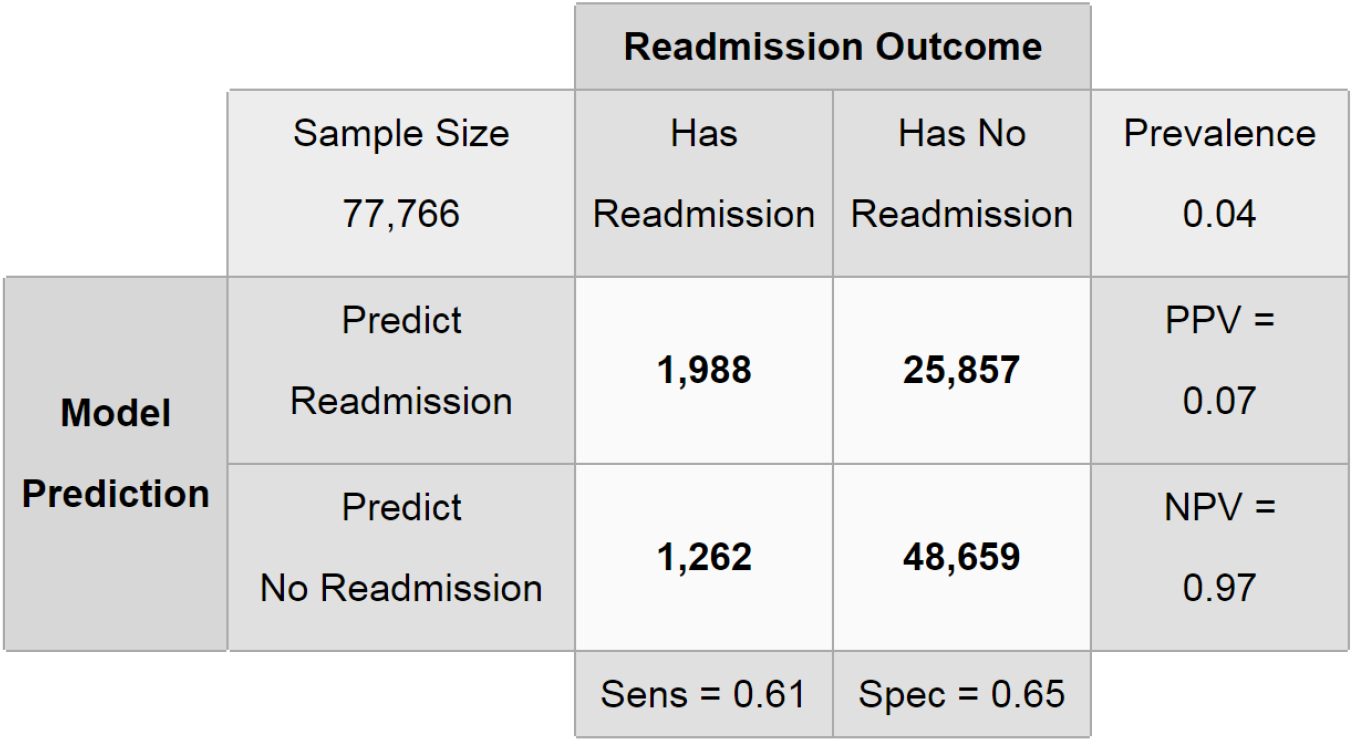
`RF` confusion matrix for 7-day unplanned readmissions on training data (OOB) using 20:1 cost ratio.

**Figure 5:**
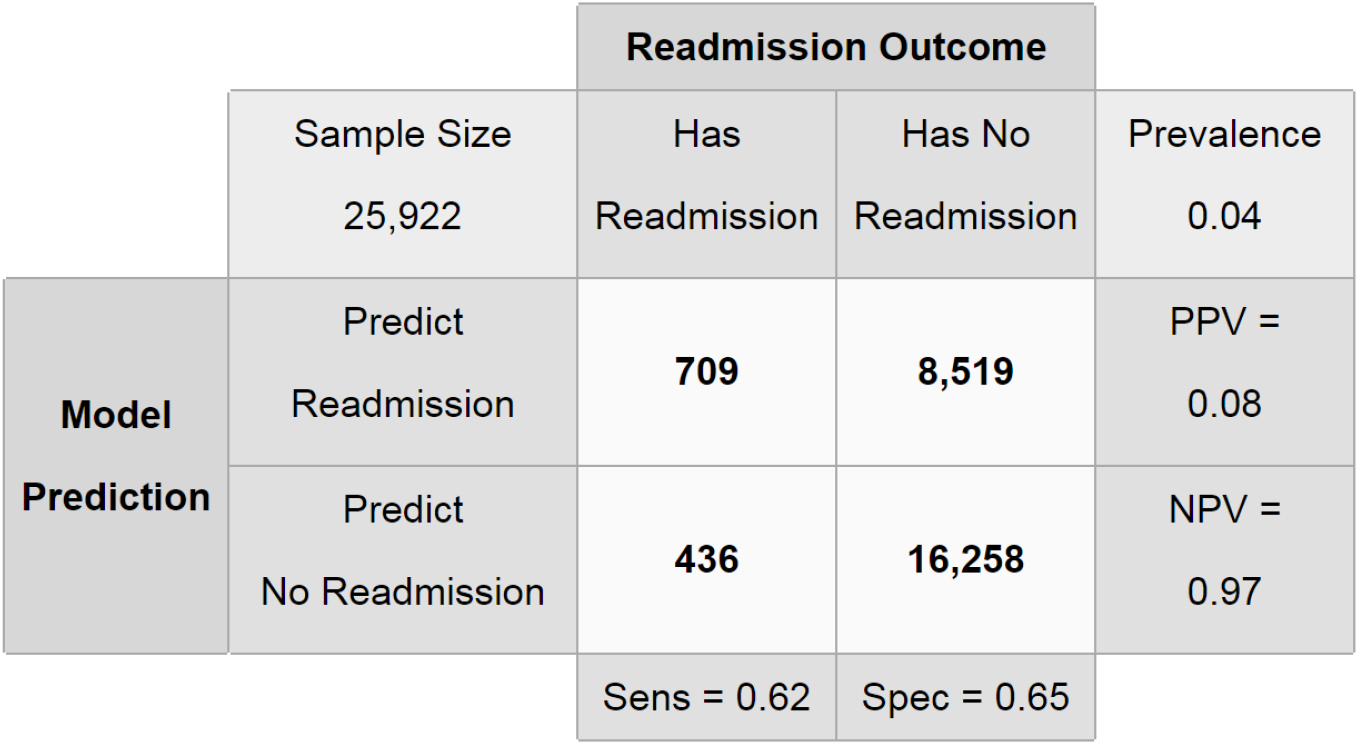
`RF` confusion matrix for 7-day unplanned readmissions on validation data using 20:1 cost ratio.

**Figure 6:**
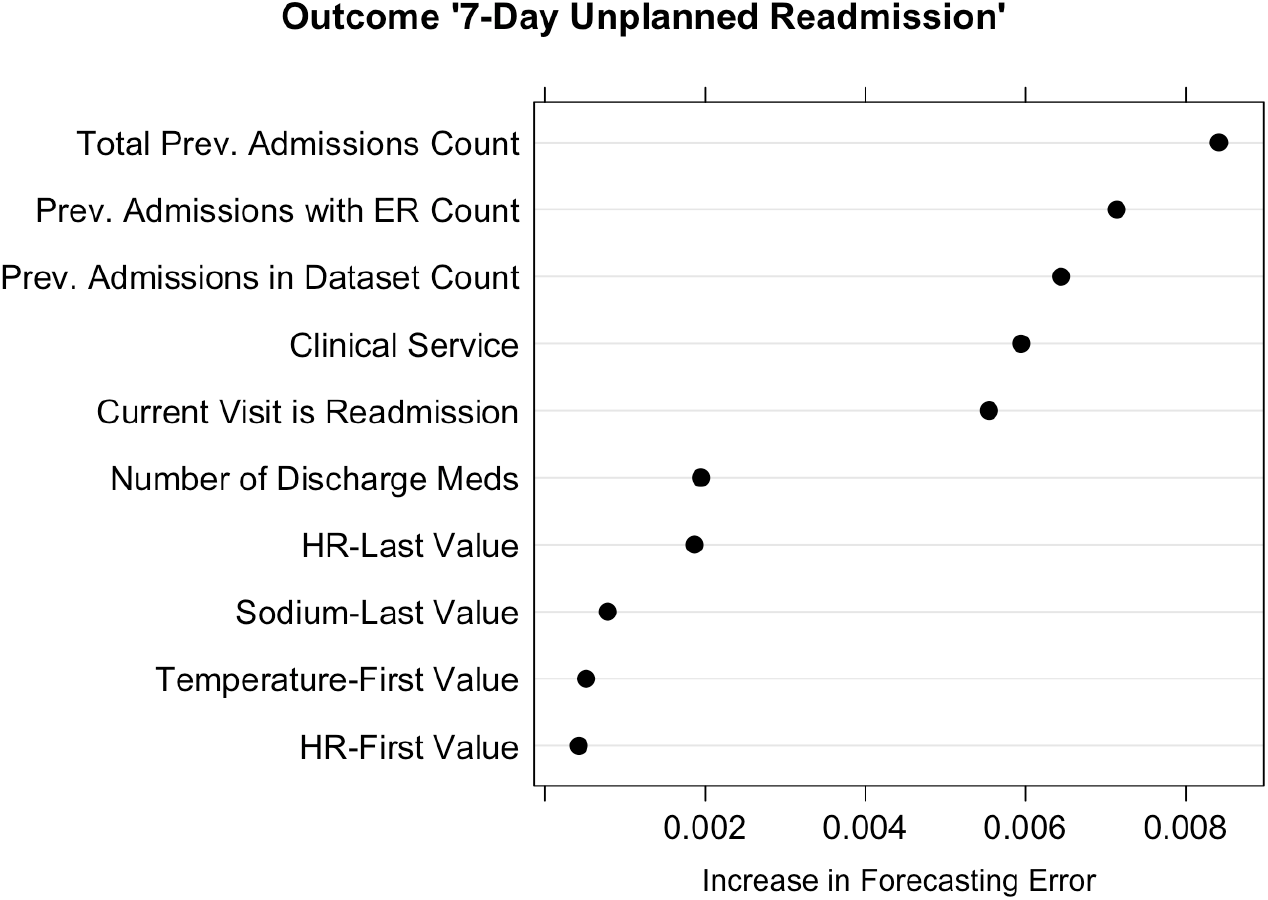
Variable importance plot for 7-day unplanned readmissions `RF` model. The top 10 variables for the class “readmission” are shown.

## 4 Discussion

### 4.1 Model performance

In this study, we report that two `RF` models sensitive to asymmetric costs can accurately predict 30-day all-cause and 7-day unplanned readmissions. We demonstrate that cost-sensitive models can achieve a balance between sensitivity and specificity, thereby overcoming the `RF` problem of majority class bias that is a hindrance to accurate prediction of a minority class, such as readmission events following discharge. We also demonstrate that the `RF` approach can utilize information encoded in EHR data to make predictions. Finally, we peek inside the black-box model and highlight the importance of variables such as health care utilization, number of discharge medications, and vital signs in the prediction of readmissions.

Considering the positive and negative predictive values for the 30-day all-cause readmissions model, when the `RF` predicts a readmission, it makes about 4 mistakes for every correct prediction. However, when it predicts no readmission, it is rarely wrong–only once in 20 predictions. The reason that the model is generally accurate when predicting no readmission, but not when predicting readmission, is a consequence of the 5:1 ratio of costs that were predefined. Given that it is of relatively high cost to misidentify a case that results in a readmission, the model casts a wide net and is more likely to predict a readmission unless it is fairly certain that the case will not result in a readmission. Health system priorities should dictate the width of this net. For example, UPHS administrators should consider the resource constraints or financial costs associated with 4 mistakes for each correct readmission prediction.

### 4.2 Comparison to Previous Results

A previous modeling effort for 30-day all-cause readmissions for UPHS produced sensitivity and specificity values of 39% and 84%, respectively, and a PPV and NPV of 30% and 89%, respectively, on prospective data [12]. We see that our `RF` approach drastically improves sensitivity with some reduction in specificity and improves the NPV with a slight reduction in the PPV. It is important to note, however, that this is not a fair comparison. The results for the previous model mathematically imply a cost ratio of roughly 1.5:1, as compared to our current cost ratio of 5:1. And an honest comparison of models under asymmetric costs requires proper tuning of all of the models under comparison to the desired cost ratio [20]. Given the lower cost of false negatives in the previous model versus ours, it is not surprising to see such a high value for specificity.

Other recent studies have used large EHR datasets to construct readmission prediction models. Shadmi et al. [33] developed a logistic regression model to predict 30-day unplanned readmissions in patients admitted to the general medicine service. They evaluated 76 candidate variables for their model and presented two potential risk thresholds. The higher threshold exhibited an effective cost ratio of 2:1 with a sensitivity of 22% and specificity of 92%, while the lower threshold had an effective for cost ratio in the validation set of 1:1.5 with a sensitivity of 37% with specificity 83%. Amarasingham et al. [34] evaluated 30 candidate variables to create a logistic regression model to predict 30-day all-cause readmissions or death in general medicine patients. The effective cost ratio in the validation set for the chosen threshold was 1:3.7 with a sensitivity of 49% and specificity of 73%.

Some readers may wonder why we are considering forecasted classes and confusion tables rather than probability outputs and c-statistics to evaluate our candidate model. To compute a c-statistic would require simply changing the threshold on the number of votes needed for classification as an unplanned readmission as is done for logistic regression. However, as previously discussed, when simply altering the threshold on the final votes, asymmetric costs are no longer reflected through the entire model construction. Also, to implement a model in practice, decisions must be made by stakeholders about the tradeoff of errors. Hence, examining the performance of the model correctly tuned to reflect stakeholder preferences should be most appropriate. A higher c-statistic value does not necessarily imply better predictive performance at a particular cost ratio, all else equal.

### 4.3 Future Directions

While the prototype models presented in this paper show promise for predicting readmissions, there is still substantial opportunity to improve the predictions. There are almost certainly useful covariates missing from the available data set that could improve predictive accuracy, and other machine learning algorithms such as support vector machines [35] and stochastic gradient boosting [30] could be explored. While `RF` shows consistently strong performance against such competitors, performance is ultimately dataset dependent. Additionally, the target outcomes could be refined to focus on preventable readmissions [25], or readmissions for specific diseases or medical conditions. After a model is constructed, providers are then presented with the challenge of integrating the prediction tool into practice, and designing and testing interventions to be used when a patient meets threshold criteria for readmission risk. Finally, the machine learning framework presented herein is highly flexible and can be applied to a broad set of health system prediction tasks, such as predicting length of stay and assessing risk of mortality.

## 5 Conclusion

In summary, we have presented a data-driven machine learning approach to predicting hospital readmissions derived from EHR data obtained at the University of Pennsylvania Health System. We employed a machine learning approach that could process a large set of patient and visit-level information to discover patterns in the data associated with readmissions. Our approach demonstrates that cost asymmetries can be integrated into the construction of `RF` models such that their predictions best reflect user needs and values. Models such as those presented herein could be implemented into an EHR so that a prediction can be produced at time of discharge for each patient. This prediction can help target scarce health system resources to the patients in need of transitional care interventions, resulting in improved patient outcomes and reduced health system costs.

## Data Availability

Aggregate data is presented in the manuscript, but our IRB approval does not currently authorize sharing of study data with those outside of the research team.

## Abbreviations

CMS: Centers for Medicare and Medicaid Services
EHR: Electronic health record
UPHS: The University of Pennsylvania Health System
RF: Random forest
OOB: Out-of-bag

## Declarations

## Acknowledgements

We would like to thank Christina Liao, Valeriy Rastorguev, Carrie Shi, Olivia Wang, Nayim Khemais, C. William Hanson, Joanne M. Resnic, Joel Betesh, Michael Draugelis, Alexander Roederer, Barry D. Fuchs, and Mark E. Mikkelsen for their input in developing the model examined in this study, their assistance in data acquisition, and for advice regarding data analysis.

## Author Contributions

JB, CAU and RB participated in the conception, coordination, and design of the study as well as data analysis and interpretation, and drafted and revised the manuscript. AH participated in the study design, data analysis and interpretation, and acquired the data. CAB participated in the study design and data analysis and interpretation, and drafted part of the manuscript. BC, AK, CAB, RG, and EC participated in study design, data analysis and interpretation, and contributed revisions to the draft manuscript. All authors read and approved the final manuscript.

## Consent for Publication

Not applicable.

## Ethics Approval and Consent to Participate

The study received expedited approval and a Health Insurance Portability and Accountability Act (HIPAA) waiver from the University of Pennsylvania Institutional Review Board.

## Appendix A: Full Variable Importance Plots

**Figure 7:**
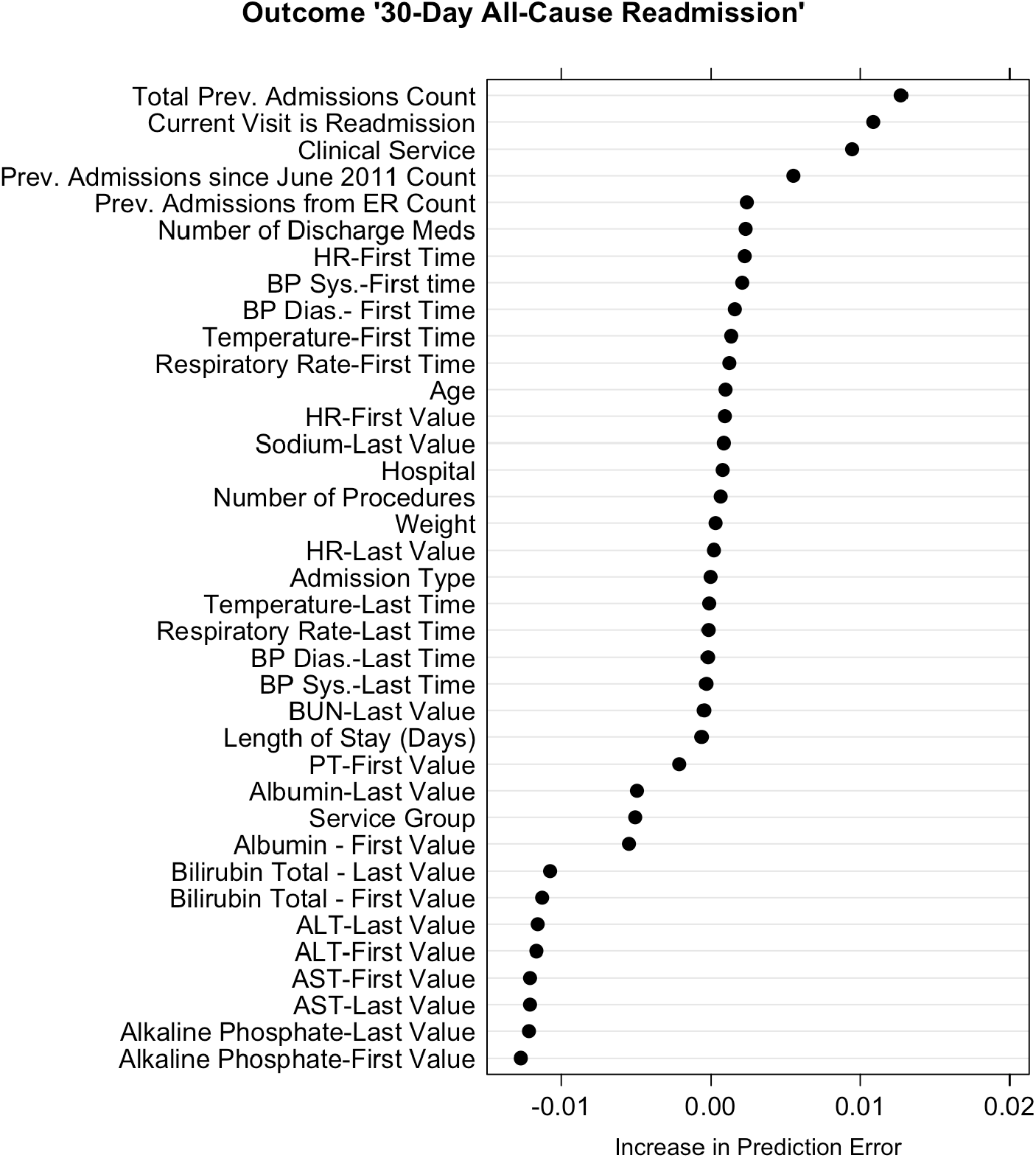
Variable importance plot for 39 predictor variables for 30-day all-cause readmissions. Lab values used in the model were the first and last recorded values during the admission or missing. To account for missingness, lab measurements (and times) were categorized into deciles with an additional category for missingness. “Service” represents the discharging service, “Service Group” is a broader grouping of discharging services, and “Admission Type” indicates whether it was a routine/elective visit or an emergency visit. Variables with negative importance scores are not necessarily useless as they may be important in interaction effects captured by the model.

**Figure 8:**
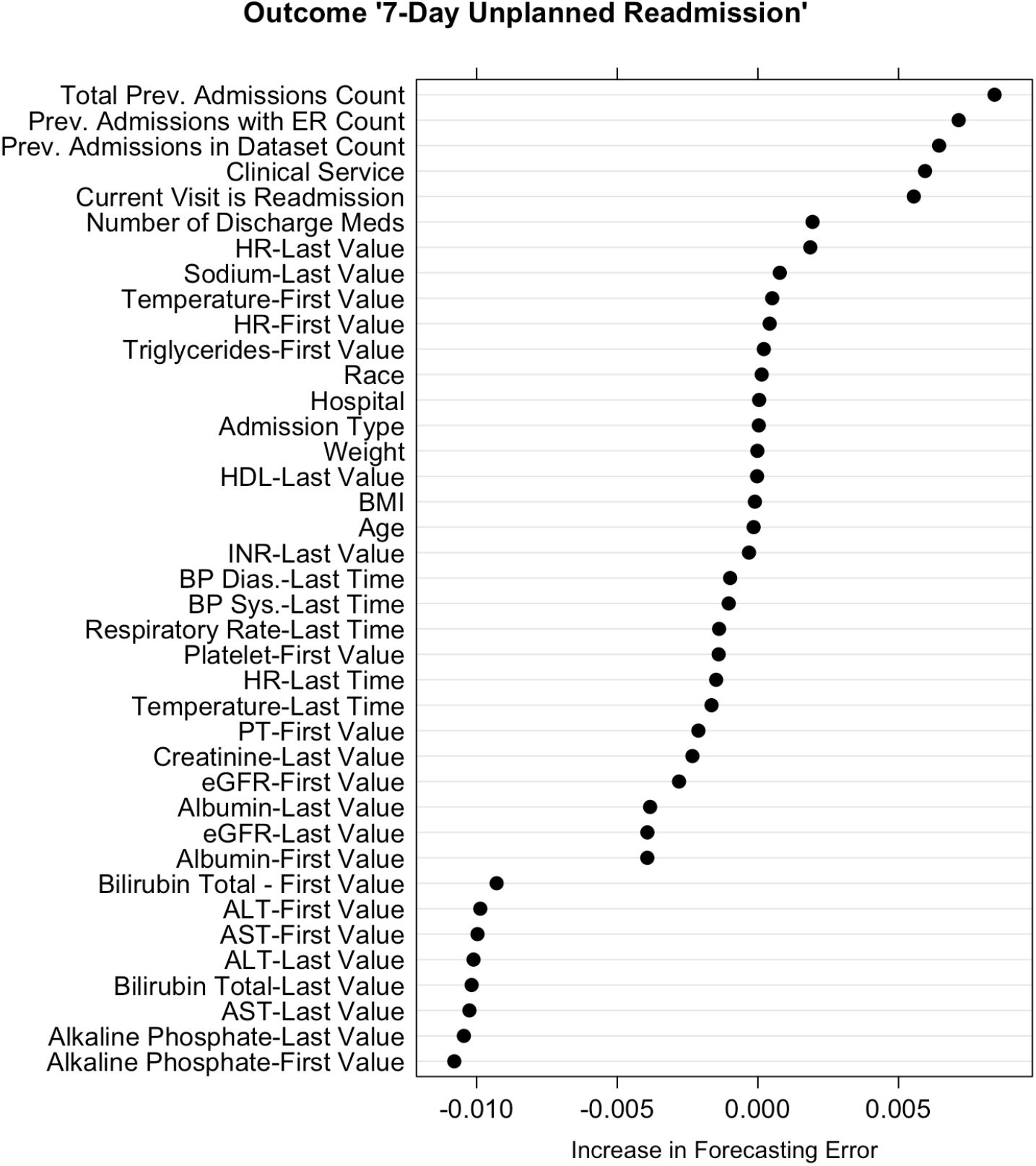
Variable importance plot for 39 predictor variables for 7-day unplanned readmissions. Lab values used in the model were the first and last recorded values during the admission or missing. To account for missingness, lab measurements (and times) were categorized into deciles with an additional category for missingness.“Service” represents the discharging service and “Admission Type” indicates whether it was a routine/elective visit or an emergency visit. Variables with negative importance scores are not necessarily useless as they may be important in interaction effects captured by the model.

## References

1. SF Jencks, MV Williams, and EA Coleman. Rehospitalizations among patients in the medicare fee-for-service program. New England Journal of Medicine, 360(14):1418–1428, 2009.

2. N Allaudeen, A Vidyarthi, J Maselli, and A Auerbach. Redefining readmission risk factors for general medicine patients. Journal of Hospital Medicine, 6(2):54–60, 2011.

3. N Bisharat, C Handler, and N Schwartz. Readmissions to medical wards: Analysis of demographic and socio-medical factors. European journal of internal medicine, 23(5):457–460, 2012.

4. N Allaudeen, JL Schnipper, EJ Orav, RM Wachter, and AR Vidyarthi. Inability of providers to predict unplanned readmissions. Journal of general internal medicine, 26(7):771–776, 2011.

5. EH Bradley, L Curry, LI Horwitz, H Sipsma, JW Thompson, M Elma, MN Walsh, and HM Krumholz. Contemporary evidence about hospital strategies for reducing 30-day readmissionsa national study. Journal of the American College of Cardiology, 60(7):607–614, 2012.

6. EH Bradley, L Curry, LI Horwitz, H Sipsma, Y Wang, MN Walsh, D Goldmann, N White, I. Piña, and HM Krumholz. Hospital strategies associated with 30-day readmission rates for patients with heart failure. Circulation: Cardiovascular Quality and Outcomes, 6(4):444–450, 2013.

7. AG Au, FA McAlister, JA Bakal, J Ezekowitz, P Kaul, and C van Walraven. Predicting the risk of unplanned readmission or death within 30 days of discharge after a heart failure hospitalization. American heart journal, 164(3):365–372, 2012.

8. EH Bradley, O Yakusheva, LI Horwitz, H Sipsma, and J Fletcher. Identifying patients at increased risk for unplanned readmission. Medical care, 51(9):761–766, 2013.

9. R Amarasingham, PC Patel, K Toto, LL Nelson, TS Swanson, BJ Moore, B Xie, S Zhang, KS Alvarez, Y Ma, et al. Allocating scarce resources in real-time to reduce heart failure readmissions: a prospective, controlled study. BMJ quality & safety, 22(12):998–1005, 2013.

10. KH Bowles, A Hanlon, D Holland, SL Potashnik, and M Topaz. Impact of discharge planning decision support on time to readmission among older adult medical patients. Professional case management, 19(1):29–38, 2014.

11. FS Ahmad, JP Metlay, FK Barg, RR Henderson, and RM Werner. Identifying hospital organizational strategies to reduce readmissions. American Journal of Medical Quality, 28(4):278–285, 2013.

12. CA. Baillie, C VanZandbergen, G Tait, A Hanish, B Leas, B French, William HC, M Behta, and CA Umscheid. The readmission risk flag: Using the electronic health record to automatically identify patients at risk for 30-day readmission. Journal of Hospital Medicine, 8(12):689–695, 2013.

13. John Billings, Ian Blunt, Adam Steventon, Theo Georghiou, Geraint Lewis, and Martin Bardsley. Development of a predictive model to identify inpatients at risk of re-admission within 30 days of discharge (parr-30). BMJ open, 2(4), 2012.

14. J Donzé, D Aujesky, D Williams, and JL Schnipper. Potentially avoidable 30-day hospital readmissions in medical patients: derivation and validation of a prediction model. JAMA internal medicine, 173(8):632–638, 2013.

15. G de Lissovoy. Big data meets the electronic medical record: A commentary on” identifying patients at increased risk for unplanned readmission”. Medical care, 51(9):759–760, 2013.

16. S Chandra, D Agarwal, A Hanson, JC Farmer, BW Pickering, O Gajic, and V Herasevich. The use of an electronic medical record based automatic calculation tool to quantify risk of unplanned readmission to the intensive care unit: A validation study. Journal of critical care, 26 (6):634–e9, 2011.

17. W Zhang, F Zeng, X Wu, X Zhang, and R Jiang. A comparative study of ensemble learning approaches in the classification of breast cancer metastasis. In Bioinformatics, Systems Biology and Intelligent Computing, 2009. IJCBS’09. International Joint Conference on, pages 242–245. IEEE, 2009.

18. SY Peng, YC Chuang, TW Kang, and KH Tseng. Random forest can predict 30-day mortality of spontaneous intracerebral hemorrhage with remarkable discrimination. European Journal of Neurology, 17(7):945–950, 2010. ISSN 1468-1331.

19. L Breiman. Random Forests. Machine learning, pages 5–32, 2001.

20. R Berk and J Bleich. Statistical procedures for forecasting criminal behavior: A comparative assessment. Criminology and Public Policy, 12: 513–544, 2013.

21. DR Cutler, TC Edwards Jr, KH Beard, A Cutler, KT Hess, J Gibson, and JJ Lawler. Random forests for classification in ecology. Ecology, 88 (11):2783–2792, 2007.

22. C Strobl, A Boulesteix, A Zeileis, and T Hothorn. Bias in random forest variable importance measures: Illustrations, sources and a solution. BMC bioinformatics, 8(1):25, 2007.

23. M Fernández-Delgado, E Cernadas, S Barro, and D Amorim. Do we need hundreds of classifiers to solve real world classification problems? The Journal of Machine Learning Research, 15(1):3133–3181, 2014.

24. KE Joynt and AK Jha. A path forward on medicare readmissions. New England Journal of Medicine, 368(13):1175–1177, 2013.

25. JG Lavenberg, B Leas, CA Umscheid, K Williams, DR Goldmann, and S Kripalani. Assessing preventability in the quest to reduce hospital readmissions. Journal of Hospital Medicine, 9(9):598–603, 2014.

26. L Breiman. Statistical Modeling: The Two Cultures. Statistical Science, 16(3):199–231, 2001.

27. A Liaw and M Wiener. Classification and regression by randomforest. R News, 2(3):18–22, 2002.

28. C Chen, A Liaw, and L Breiman. Using random forest to learn imbalanced data. Technical report, Department of Statistics, UC Berkeley, 2004. URL http://statistics.berkeley.edu/sites/default/files/tech-reports/666.pdf.

29. T Hastie, R Tibshirani, and JH Friedman. The Elements of Statistical Learning. Springer Science, second edition, 2009.

30. JH Friedman. Stochastic gradient boosting. Computational Statistics & Data Analysis, 38(4):367–378, 2002.

31. HA Chipman, EI George, and RE McCulloch. Bart: Bayesian additive regression trees. The Annals of Applied Statistics, pages 266–298, 2010.

32. R Díaz-Uriarte and S Alvarez de Andrés. Gene selection and classification of microarray data using random forest. BMC bioinformatics, 7:3, January 2006.

33. E Shadmi, N Flaks-Manov, M Hoshen, O Goldman, H Bitterman, and RD Balicer. Predicting 30-day readmissions with preadmission electronic health record data. Medical care, 53(3):283–289, 2015.

34. R Amarasingham, F Velasco, B Xie, C Clark, Y Ma, S Zhang, D Bhat, B Lucena, M Huesch, and EA Halm. Electronic medical record-based multicondition models to predict the risk of 30 day readmission or death among adult medicine patients: validation and comparison to existing models. BMC medical informatics and decision making, 15(1):39, 2015.

35. VN Vapnik. Statistical learning theory, volume 2. Wiley New York, 1998.

